# Forms of support and experiencing maltreatment and disrespect during childbirth at a health facility: a self-reported cross-sectional study in Ghana

**DOI:** 10.1101/2022.07.19.22277809

**Authors:** Agnes Asare, Philip Teg-Nefaah Tabong

## Abstract

Providing support to women and encouraging women to deliver in health facility settings (skilled delivery) is one strategy to improve maternal health outcomes in developing countries. However, fear of maltreatment and disrespect during labour and delivery, have been reported as barriers to facility delivery. This study was therefore conducted to assess postnatal women’s self-reported experience and type of maltreatment and disrespect during delivery. Using a cross sectional study design, 313 women were randomly selected from the three health facilities located in the La-Nkwantenang-Madina Municipality in the Greater Accra region. The data was analysed using STATA 15. Bivariable data analysis using Pearson’s Chi-square was used to determine the association between socio-demographic characteristics, reproductive history and experience of maltreatment. Logistic regression analysis was done to determine the strengths of association. More than half (54.3%) of the postnatal women were encouraged to have support persons during labour and delivery. About 75.7% reported experiencing some form of maltreatment and disrespect. About a fifth (19.8%, n=62) of the women experienced physical abuse, 9.3% (n=29) experienced undignified care, 7.7% (n=24) of the women were detained or confined against their will, and 60.4% (n=189) indicated they did not receive consented care. The study concludes that maltreatment and disrespect during labour is widespread. Expanding health facilities may not achieve desired skilled or facility-based delivery without improving the birth experience of women.

## Introduction

Maternal mortality remains in challenge globally. The majority (99%) of these avoidable deaths occur in low-income countries [1]. Maternal death has disastrous outcomes for families, communities and country with significant economic effect resulting in broken homes and motherless children [2]. The crucial hurdle to the utilization of health services for women that contributes to maternal death is a delay in the decision to seek care, delay in arrival at a health facility and lack of provision of satisfactory care [3]. The lack of provision and adequate care buttress the need to improve women’s access to maternity care and promote birth in health facilities with skilled attendants [4].

According to Souza et al., the majority of maternal deaths have been found to be avoidable [5]. Maternal and neonatal health can improve and complications avoided when women access skilled respectful care during pregnancy and childbirth from health care providers [6]. Women’s experiences and their perceptions of maternity care are formed principally on client-provider interpersonal relationship [7]. The major drawback to accessing skilled care for routine and complicated births is poor interactions between the patient and the health provider [8,9]. Furthermore, the Respectful Maternity Care Charter as published by White Ribbons Alliance recognizes that women’s perceptions to childbirth are an essential component of quality health care and their autonomy, dignity, feeling, choices and preferences must be respected [10].

The Universal Rights on Childbearing Women, is enshrined with international human rights and comprises of right to information, informed consent, privacy and confidentiality, dignity and respect, and free from physical abuse and discrimination. The care received during childbirth must emphasize these and not solely on the prevention of death of mother and baby [11]. Disrespect and abuse during pregnancy and childbirth have been identified as an important indicator of poor quality care and a barrier to improved maternal outcomes with inadequate data on the scope and magnitude, especially in urban areas of low-income countries [12].

According to Bohren et al., the facilitator and barrier to future facility-based delivery are dependent on experience during child birth and pregnancy outcomes [3]. Also, women who experience negative interactions in a facility are reluctant to report back for subsequent deliveries [13]. According to Okafor the high rates and contribution of disrespect and abuse continue to favour home deliveries with no skilled attendance in Nigeria [14]. This can lead to poor maternal and neonatal health outcomes and encourage the use of traditional birth attendants. In Earlier study in rural community in Ghana reported that women experience maltreatment during delivery which tend to favour the use traditional birth attendants (TBA) [15]. Such an encounter can be a barrier to future facility-based delivery in a setting where women view TBAs as more dependable than health facility personnel, as providing high-quality care in terms of supportive care, skill and emotional help during childbirth [3].

At La-Nkantanang Madina Municipality in Accra where the study was conducted, supervised deliveries have marginally improved from 45.3% to 62.8% for the first half of the year from 2012 to 2017. However, this is below 92.1% reported in the Greater Accra in a nationwide survey [16]. Given the reported correlate between maltreatment and accessing skilled delivery, this study was conducted to document self-reported experiences of maltreatment.

## Methods and materials

### Ethical approval

This study protocol was reviewed and approved by the Ghana Health Service Ethics Review Committee (GHS-ERC 020/03/18). All participants signed a written consent form before participating in the study.

### Study design

A quantitative cross-sectional study was conducted at three selected health facilities in La-Nkwantenang-Madina Municipality (LNMM) in the Greater Accra region of Ghana. The choice of study was informed by an earlier study in South West Ethiopia [17].

### Study settings

The study was conducted at the three facilities mentioned earlier in La-Nkwantanang Madina Municipality; Pentecost Hospital, Madina Polyclinic at Kekeli and Madina Polyclinic at Rawling circle. These facilities render maternal and child health care to residents of the Municipality and beyond.

### Study Population

This study was conducted among postnatal women within the reproductive ages of 15-49 years who delivered at these selected facilities and were attending postnatal clinic at the time of the study.

### Inclusion and exclusion Criteria

Women who have delivered either vaginally or by emergency caesarean section, reporting for their sixth-week postnatal review were included in the study. Those who had a vaginal birth or emergency caesarean section after induction of labour as well as mothers in critical medical condition or experienced stillbirths but attended postnatal for examination were excluded from the study. Women who had experienced stillbirth were excluded because they may be going through emotional distress which made it unethical to interview them.

### Sample Size Determination

A single population proportion formula was used to estimate the sample size with an assumption of 5% precision and 95% confidence. An assumption that 72% of labouring mothers would face at least one form of disrespect and abuse during childbirth was used based on earlier study [18]. Based on this, a minimum sample size of 309 was determined. In studies using postnatal women, it has been reported that a non-response rate of 2-10% should be expected. However, during the pilot study, no non-response was recorded. Nonetheless, the minimum response rate was adopted as advised. This, therefore, increased the sample size to 316.

### Sampling Technique

The allocation of the sample to the three health facilities was made proportionately, based on the number of clients who received childbirth services at each facility in the month preceding the data collection period. The average number of deliveries per month in Pentecost Hospital, Madina Polyclinic (RC) and Madina Polyclinic Kekele were 310,40 and 108 respectively (Table 1).

**Table 1:** Proportionate Allocation to Health Facilities

| <u>Health Facility</u> | <u>Population (N)</u> | <u>Population proportion</u><br>$P = \frac{N}{458} * 100$ | <u>Number selected</u><br>$N = (p * 316)$ |
| --- | --- | --- | --- |
| Pentecost Hospital<br>Madina (PHM) | 310 | 0.68 | 215 |
| Madina Polyclinic<br>(RC) | 40 | 0.09 | 28 |
| Madina Polyclinic<br>Kekele (MPK) | 108 | 0.23 | 73 |
| Total | 458 | 1.00 | 316 |

The postnatal clinic runs every week hence the sample size of mothers to be interviewed per week for Pentecost Hospital, Madina Polyclinic (RC) Madina Polyclinic Kekele were 36, 5, and 13 respectively. A simple random sampling without replacement was conducted at the selected facility to obtain the participants for the study. At each facility, a list of mothers attending postnatal clinic per week was obtained from the medical records and assigned numbers from one to the last number. Each number was written on a paper of the same size and put in a bowl with a blindfolded person to pick 36, 5, and 13 papers at Pentecost Hospital, Madina Polyclinic (RC) Madina Polyclinic Kekele respectively. The mothers whose names corresponded to the numbers selected at each facility were recruited into the study. If a client declined to participate, a replacement was done.

### Study variables

Disrespect has seven categories: (1). Physical abuse: slapping/hitting or physical force on the mother; (2). Non-dignified care: mother experience rudeness, scolding or insults; (3). Non-consented care: No informed consent or information dissemination; (4). No privacy: curtains or other visual barriers not use; (5). Discrimination: mother prejudiced on grounds of clinic state; (6). Neglect: mother was left alone or unattended to; (7). Detention: mother delayed in health facility against her will.

A woman was considered to have received disrespectful care if she experienced at least one of the seven categories of disrespect and abuse. For a specific category of abuse and disrespect with more than one verification criterion, a woman was labelled “abused and disrespected” in that category if she was abused and disrespected in at least one of the verification criteria during childbirth. The rest of the variables were socio-demographic such as age, marital status, education, religion, parity, mode of delivery

### Data collection and Tools

A self-administered structured questionnaire was used for data elicitation. Participants who could not read, the Research Assistants interpreted the questions and helped them filled the questionnaire. The questionnaires were administered on a one-on-one basis, to the participants. The first part of the questionnaire elicited participants socio-demographic characteristics/ client-related factors. The second part assessed disrespectful care and during labour and childbirth. The tool was adapted from the Maternal and Child Health Integrated Program (MCHIP) tool kit instrument that assesses and improves respectful maternity care [19]. An approach centred on the individual, touches on respect for beliefs, traditions and culture; the right to information and privacy; confidentiality; consent and preference in care; choice of a companion during labour and birth; freedom of movement during labour; non-separation of mother and newborn and prevention of institutional violence, including abusive and disrespectful care. The last part of the questionnaire assessed the preference for facility-based delivery in subsequent births based on their experience with their last birth. It took on an average of 40 minutes to complete or administer a questionnaire.

### Data Processing and Analysis

Data was entered, cleaned and analysed using STATA software version15. Frequency and percentages were used to describe the demographic characteristics of the study participants. The form of disrespect and abuse were also summarized descriptively using the frequency and percentages. The Pearson’s chi-square test of association was used to assess socio-demographic factors and experiencing at least one of the forms of disrespectful. The multivariable logistic regression model was used to estimate the crude and adjusted odds ratio of the factors significantly associated with the experience of at least one form of disrespectful care from Pearson’s chi-square test. All statistical tests with p-values below 0.05 were considered significant and a 95% confidence interval.

## Results

### Socio-demographic characteristics of respondents

A of 316 questionnaire were administered, however 313 were retrieved (response rate of 99%). About half (49.8%, n=156) were within the age range 25 to 34 years, 22.0% (n=69) were single whilst 43.8% (n=137) were married, 17.9% (n=56) had no formal education whilst 12.5% (n=39) had tertiary level of education, 22.4% (n=70) were unemployed. Most (68.7%, n=215) of the women delivered at PHM. (Table 2).

**Table 2:** Background characteristics of Postnatal women

| **Socio-demographic characteristics and reproductive history** | **Frequency**<br>**(n=313)** | **Percentage**<br>**(%)** |
| --- | --- | --- |
| **Age group** |  |  |
| <25 | 111 | 35.46 |
| 25-34 | 156 | 49.84 |
| 35-44 | 46 | 14.70 |
| **Marital status** |  |  |
| Single | 69 | 22.04 |
| Married | 137 | 43.77 |
| Co-habiting | 81 | 25.88 |
| Divorced/Widowed | 26 | 8.31 |
| **Highest education** |  |  |
| No education | 56 | 17.89 |
| Primary | 107 | 34.19 |
| Secondary | 111 | 35.46 |
| Tertiary | 39 | 12.46 |
| **Employment** |  |  |
| Unemployed | 70 | 22.36 |
| Civil servant | 44 | 14.06 |
| Traders | 146 | 46.65 |
| Others | 53 | 16.93 |
| **Income (GHS)** |  |  |
| None | 18 | 5.75 |
| <500 | 113 | 36.10 |
| 500-1000 | 88 | 28.12 |
| >1000 | 94 | 30.03 |
| **Religion** |  |  |
| Muslim | 115 | 36.74 |
| Christians | 198 | 63.26 |
| **Parity** |  |  |
| P1-2 | 229 | 73.16 |
| P3-4 | 71 | 22.68 |
| P4+ | 13 | 4.15 |
| **Mode of delivery** |  |  |
| Emergency Caesarean Section (Em. CS) | 68 | 21.73 |
| Spontaneous Vagina Delivery (SVD) | 245 | 78.27 |
| **Facility of delivery** |  |  |
| MPK | 70 | 22.36 |
| MPR/C | 28 | 8.95 |
| PHM | 215 | 68.69 |

### Forms of Support received during labour and childbirth

Over half (54.3%, n=170) of the women were encouraged to have a support person present during delivery whilst 76.7% (n=240) liked the idea of support persons being present. A third (33.2%, n=104) had their mothers as labour companions, 22.4% (n=70) had their husband or partners present whilst 10 (3.2%) had no one present during delivery. Less than half 34.5% (n=108) of the women had the freedom of movement during labour, 44.7% (n=140) were encouraged to eat foods, 50.2% (n=157) were encouraged to take fluids, 51.4% (n=161) had labour analgesia, 29.4% (n=92) had episiotomy done whilst 80.2% (n=251) had their babies given to them immediately after birth (Table 3).

**Table 3:** Forms of Support received during labour and childbirth

| Forms of support | Frequency (n) | Percentage (%) |
| --- | --- | --- |
| **Woman encouraged to have support person** |  |  |
| No | 143 | 45.69 |
| Yes | 170 | 54.31 |
| **A woman likes the idea of a support person** |  |  |
| No | 73 | 23.32 |
| Yes | 240 | 76.68 |
| **Labour companion** |  |  |
| Mother | 104 | 33.23 |
| Husband/partner | 70 | 22.36 |
| Friend | 31 | 9.90 |
| Others | 98 | 31.31 |
| None | 10 | 3.19 |
| **The woman had freedom of movement during labour** |  |  |
| No | 151 | 48.24 |
| Yes | 108 | 34.50 |
| Don't know | 54 | 17.25 |
| **Encouraged to eat food** |  |  |
| No | 173 | 55.27 |
| Yes | 140 | 44.73 |
| **Encouraged to take fluids** |  |  |
| No | 156 | 49.84 |
| Yes | 157 | 50.16 |
| **Labour analgesia** |  |  |
| No | 152 | 48.56 |
| Yes | 161 | 51.44 |
| **Episiotomy/laceration** |  |  |
| No | 221 | 70.61 |
| Yes | 92 | 29.39 |
| **The baby was given to the mother immediately after birth** |  |  |
| No | 62 | 19.81 |
| Yes | 251 | 80.19 |

### Self-reported experience and forms of disrespect during labour and childbirth

Figure 1 shows the frequency distribution of the number of women who experienced the various forms of disrespect during the delivery at the health facilities. About a fifth (19.8%, n=62) of the women experienced physical abuse and 9.3% (n=29) were discriminated against. 38.9% (n=122) of the women experienced undignified care, 7.7% (n=24) of the women were detained or confined against their will, 60.4% (n=189) did not receive consented care, the privacy or the confidentiality of 109 (34.8%) women were not protected and 17.9% (n=56) of the women were left unattended to when they needed care.

**Figure 1:**
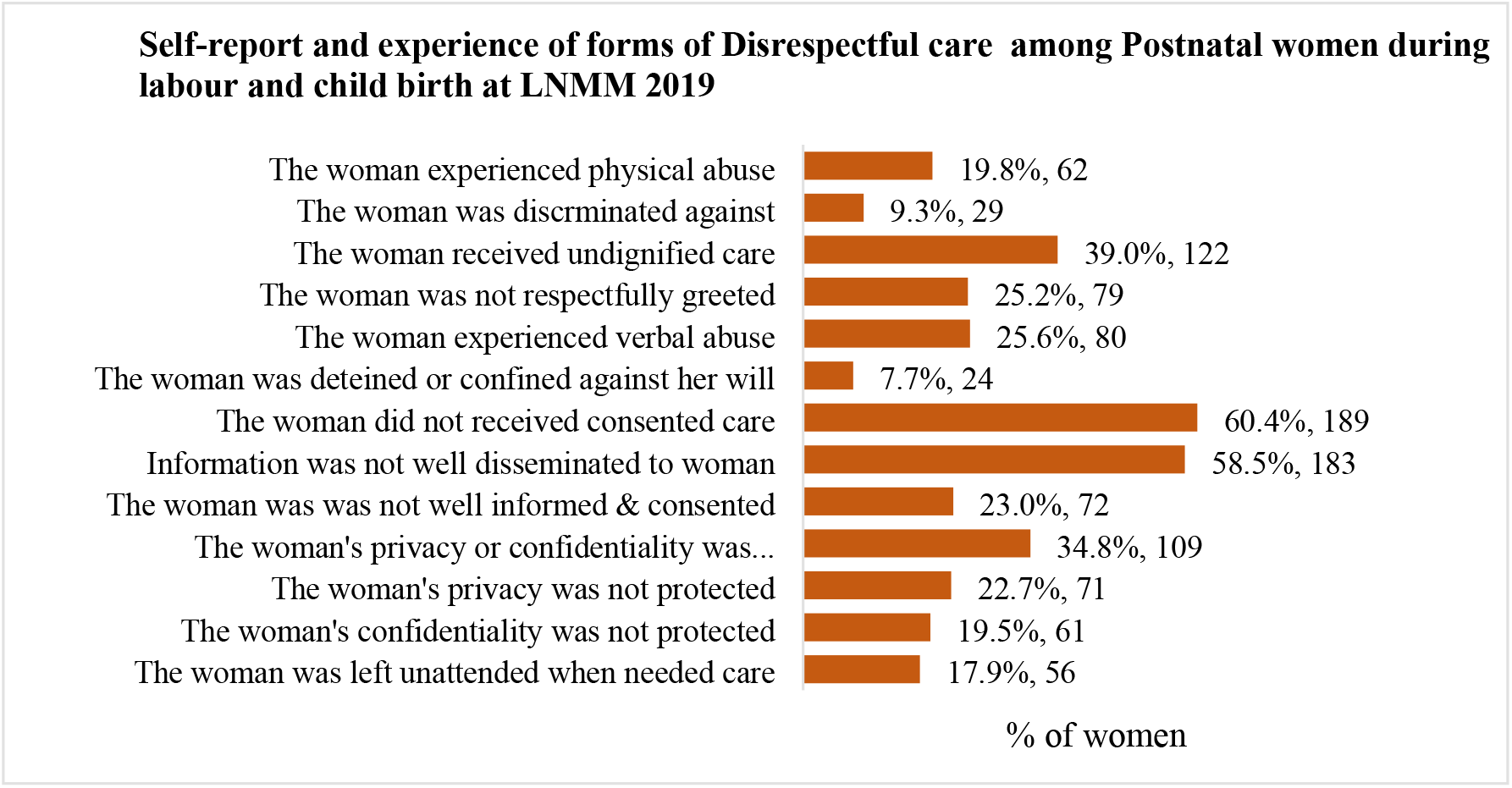
Self-reported and experience and forms of disrespect experienced during labour and childbirth

In overall, 75.7% (n=237) of the women experienced at least one form of disrespectful care during their delivery at the health facility. (Figure 2)

**Figure 2:**
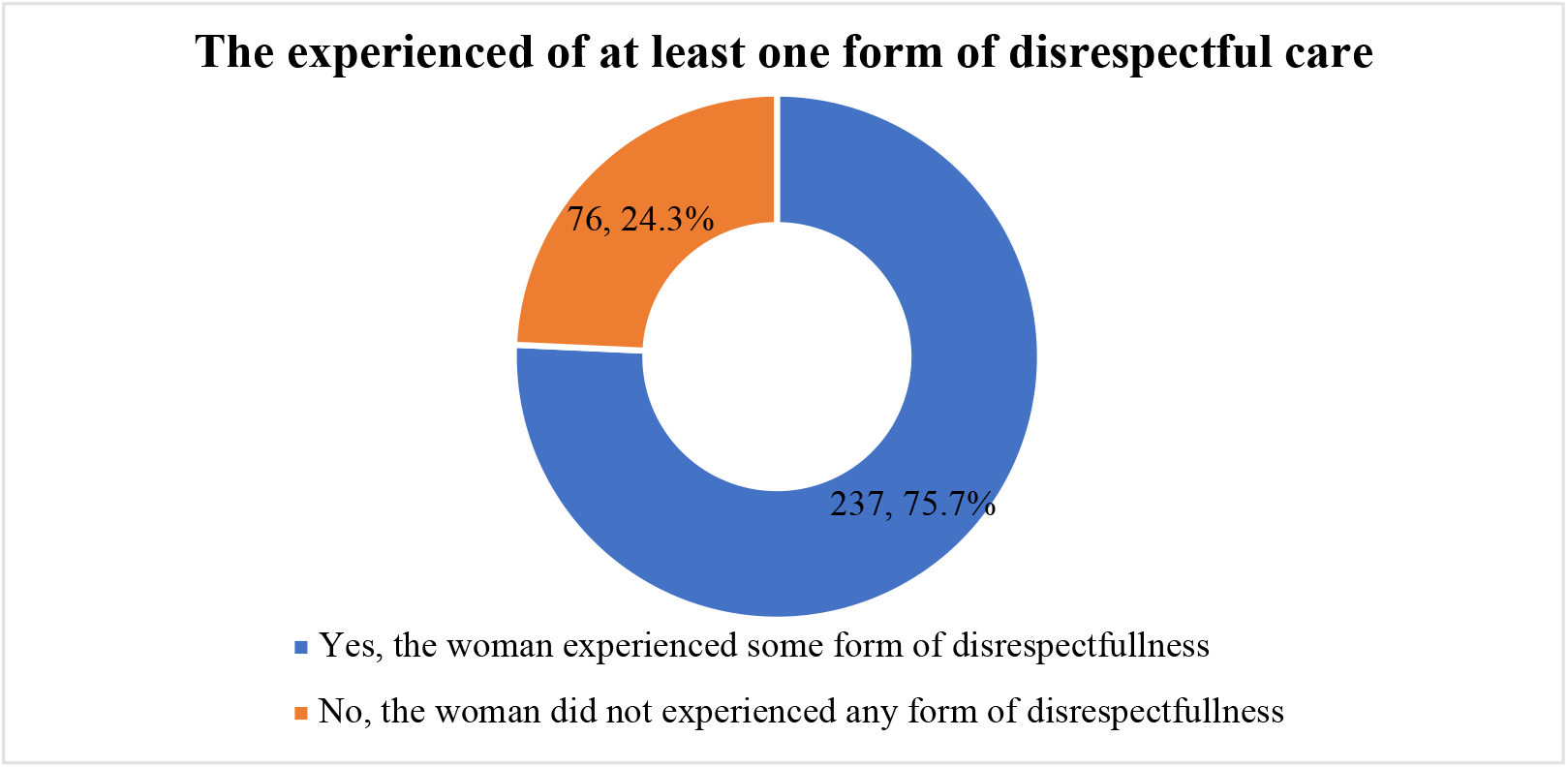
Experience of disrespectful care among Postnatal women during labour and delivery.

### Association between socio-demographic characteristics (SCD)/reproductive factors (RH) and experience of disrespectful care

The experience of at least one form of disrespectful care was 85.5% (59/69) among single women, 64.9% (89/137) among married women, 80.3% (65/81) among cohabiting women, and 92.3% (24/26) among divorced or widowed women. There was significant association between marital status and the experience of at least one form of disrespectful care (χ^2^=17.01, p=0.001).

Experience of some form of disrespectful care was higher among women who visited PHM (86.5%, 186/215) compared to those who visited MPK (58.6%, 41/70) and MPR/C (35.7%, 10/28). There was a significant association between health facility delivered at and the experience of at least one form of disrespectful care (χ^2^=49.19, p<0.001). Highest level of education (χ^2^=15.05, p=0.002), employment (χ^2^=10.97, p=0.012), religion (χ^2^=10.63, p=0.001) and mode of delivery (χ^2^=13.54, p<0.001) were also significantly associated with the experience of at least one form of disrespectful care. The woman is encouraged to have a support person (χ2=17.31, p<0.001), the woman liking the idea of a support person (χ2=24.03, p<0.001), the labour companion present (χ2=13.56, p=0.009), having the freedom of movement during labour (χ2=23.92, p<0.001), encouraged to eat food (χ2=22.79, p<0.001), encouraged to take fluids (χ2=15.39, p<0.001), having labour analgesia (χ2=20.32, p<0.001) and the baby given to mother immediately after delivery (χ2=11.06, p=0.001) were the forms of support significantly associated with the experience of at least one form of disrespectful care. (Table 4).

**Table 4:**
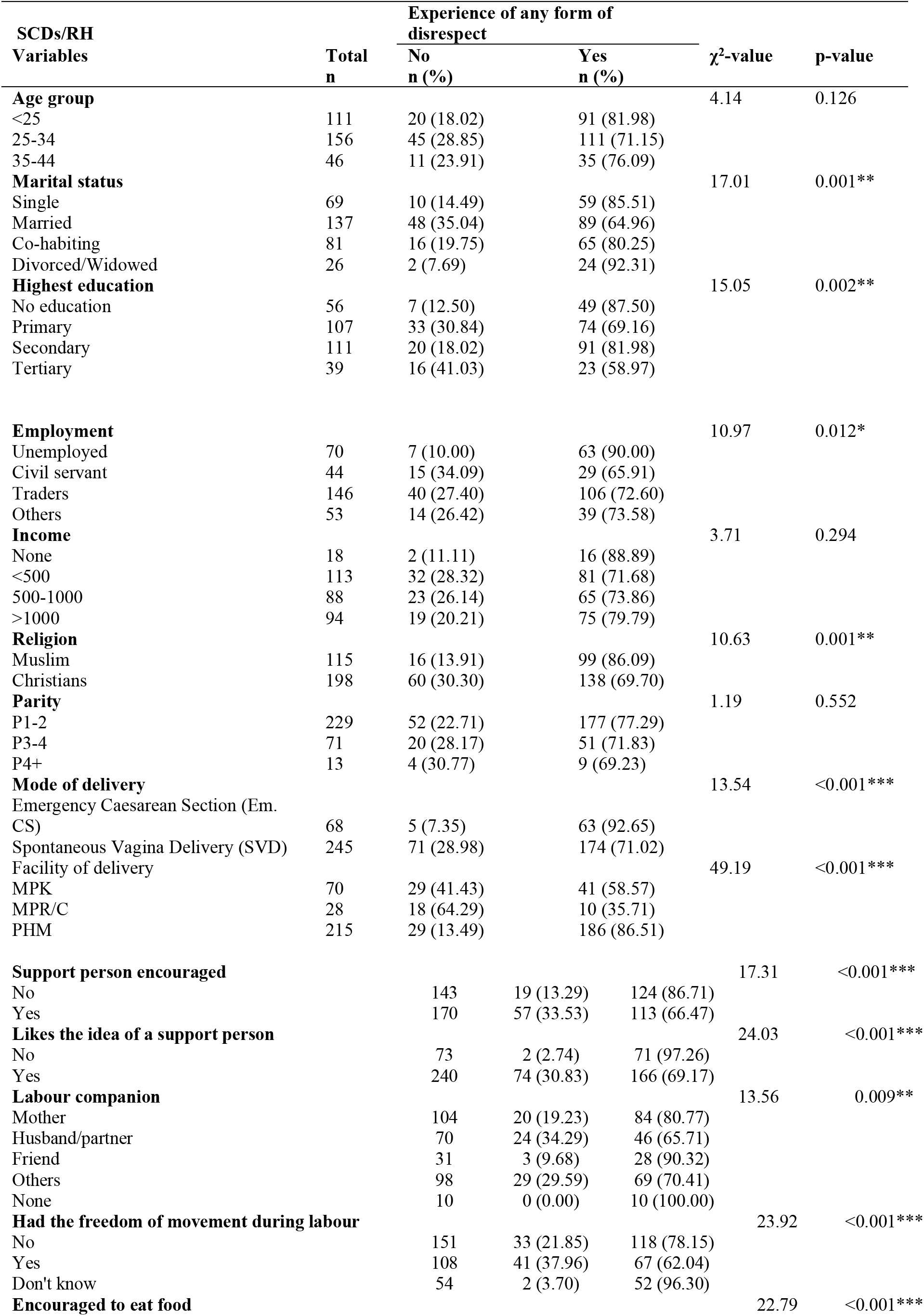
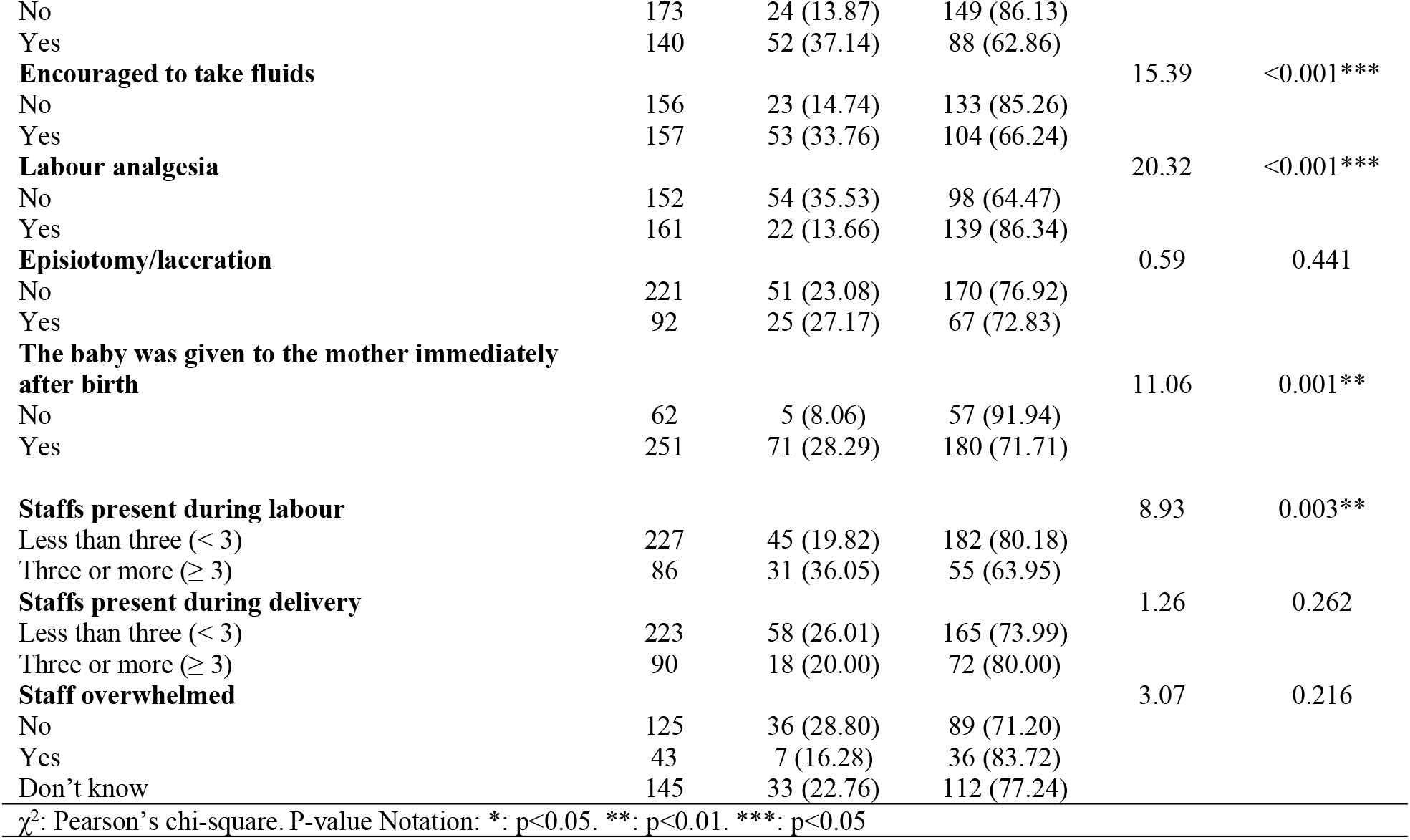
Association between Socio-demographic characteristics (SCDs)/reproductive/form of support history (RH) and experience of disrespectful care

### Correlates of the experience of disrespect during labour and delivery

Table 5 shows a multivariable analysis of the factors associated with the experience of at least one form of disrespectful care towards women delivering at the health care facility. The simple logistic regression model was used to estimate the unadjusted odds ratio whilst the multiple logistic regression model was used to estimate the adjusted odds ratio. From the adjusted logistic regression model, the odds of a woman experiencing at least one form of disrespectful care was 5 times higher for those visiting PHM compared to those visiting MPR/C (AOR: 5.01, 95% CI: 1.18-21.35, p=0.029). The odds of experiencing at least one form of disrespectful care were 2.5 times among women who were not encouraged to have a support person present compared to women who were encouraged to have a support person present during delivery (AOR: 2.48, 95% CI: 1.12-5.49, p=0.026). Also, women who did not like the idea of support person present during labour had 5-times increased odds of experiencing at least one form of disrespectful care compared to those who did not like the idea of support person present (AOR: 5.38, 95% CI: 1.08-26.81, p=0.040). (Table 5)

**Table 5:** Multivariate analysis of factors influencing the experience of any form of disrespect towards Postnatal women during labour and delivery

| **Any form of disrespect** | **Unadjusted logistic regression** |  | **Adjusted logistic regression** |  |
| --- | --- | --- | --- | --- |
| **Variables** | **UOR [95% CI]** | **P-value** | **AOR [95% CI]** | **P-value** |
| **Marital status** |  |  |  |  |
| Single | 1.00 [reference] |  | 1.00 [reference] |  |
| Married | 0.31 [0.15-0.67] | 0.003** | 1.10 [0.39-3.06] | 0.857 |
| Co-habiting | 0.69 [0.29-1.64] | 0.398 | 1.90 [0.60-5.96] | 0.272 |
| Divorced/Widowed | 2.03 [0.41-9.98] | 0.382 | 2.91 [0.50-17.02] | 0.236 |
| **Highest education** |  |  |  |  |
| No education | 4.87 [1.76-13.46] | 0.002** | 3.70 [0.78-17.64] | 0.101 |
| Primary | 1.56 [0.73-3.33] | 0.251 | 1.97 [0.55-6.97] | 0.295 |
| Secondary | 3.17 [1.42-7.05] | 0.005** | 2.72 [0.81-9.16] | 0.107 |
| Tertiary | 1.00 [reference] |  | 1.00 [reference] |  |
| **Employment** |  |  |  |  |
| Unemployed | 1.00 [reference] |  | 1.00 [reference] |  |
| Civil servant | 0.21 [0.08-0.58] | 0.003** | 0.50 [0.12-2.17] | 0.354 |
| Traders | 0.29 [0.12-0.70] | 0.005** | 0.65 [0.22-1.94] | 0.434 |
| Others | 0.31 [0.11-0.83] | 0.020* | 1.16 [0.31-4.35] | 0.830 |
| **Religion** |  |  |  |  |
| Muslim | 1.00 [reference] |  | 1.00 [reference] |  |
| Christians | 0.37 [0.20-0.68] | 0.001** | 0.88 [0.38-2.02] | 0.764 |
| **Mode of delivery** |  |  |  |  |
| Emergency Caesarean Section | 1.00 [reference] |  | 1.00 [reference] |  |
| Spontaneous Vagina Delivery | 0.19 [0.08-0.50] | 0.001** | 0.63 [0.17-2.33] | 0.486 |
| **Facility of delivery** |  |  |  |  |
| MPK | 2.54 [1.03-6.31] | 0.044* | 1.36 [0.42-4.41] | 0.611 |
| MPR/C | 1.00 [reference] |  | 1.00 [reference] |  |
| PHM | 11.54 [4.85-27.46] | <0.001*** | 5.01 [1.18-21.35] | 0.029* |
| **Woman encouraged to have support person** |  |  |  |  |
| No | 3.29 [1.85-5.87] | <0.001*** | 2.48 [1.12-5.49] | 0.026* |
| Yes | 1.00 [reference] |  | 1.00 [reference] |  |
| **A woman likes the idea of a support person** |  |  |  |  |
| No | 15.83 [3.78-66.24] | <0.001*** | 5.38 [1.08-26.81] | 0.040* |
| Yes | 1.00 [reference] |  | 1.00 [reference] |  |
| **Labour companion** |  |  |  |  |
| Mother | 1.00 [reference] |  | 1.00 [reference] |  |
| Husband/partner | 0.46 [0.23-0.91] | 0.027* | 1.28 [0.45-3.65] | 0.645 |
| Friend | 2.22 [0.61-8.05] | 0.224 | 1.12 [0.22-5.60] | 0.891 |
| Others | 0.57 [0.29-1.09] | 0.088 | 0.72 [0.30-1.76] | 0.475 |
| **The woman had freedom of movement during labour** |  |  |  |  |
| No | 1.00 [reference] |  | 1.00 [reference] |  |
| Yes | 0.46 [0.26-0.79] | 0.005** | 0.84 [0.39-1.79] | 0.642 |
| Don't know | 7.27 [1.68-31.44] | 0.008** | 3.66 [0.70-19.30] | 0.126 |
| **Encouraged to eat food** |  |  |  |  |
| No | 1.00 [reference] |  | 1.00 [reference] |  |
| Yes | 0.27 [0.16-0.47] | <0.001*** | 0.68 [0.24-1.91] | 0.467 |
| **Encouraged to take fluids** |  |  |  |  |
| No | 1.00 [reference] |  | 1.00 [reference] |  |
| Yes | 0.34 [0.20-0.59] | <0.001*** | 0.88 [0.31-2.46] | 0.806 |
| **Labour analgesia** |  |  |  |  |
| No | 1.00 [reference] |  | 1.00 [reference] |  |
| Yes | 3.48 [1.99-6.09] | <0.001*** | 2.05 [0.82-5.10] | 0.123 |

## Discussion

### Forms of support received during labour and childbirth at LNMM 2019

More than half (54.3%) of the postnatal women were encouraged to have support persons during labour and delivery. This may be as a result of inadequate space at the labour wards to allow effective execution of labour companion at these facilities or the value of the labour support underestimated. More than two-thirds (76.7%) of the women confirmed their preference for the idea of a support person during labour and delivery. This affirms the assertion that women like to be supported during labour [20]. The companions may provide psychological support to endure labour and childbirth, but also provide logistics and assistance to the staff where necessary [21]. One out of every four (22.4%) labour companions were husbands or partners which reveals a low male involvement during labour and childbirth while their mothers formed a third (33.2%) of the support persons. About a third (34.5%) of the postnatal women had freedom of movement during labour which is not encouraging. This could be as a result of either the respectful maternity care guidelines are not complied or inadequate space available at the labour ward discouraged the practice of unrestricted movement during labour.

About half of the women were encouraged to drink fluids (50.2%) and eat food (44.7%) during labour. This is also not encouraging because the process of childbirth may lead to maternal exhaustion especially if the labouring women are denied this support. The woman starved and or dehydrated, may lack the energy to expel the baby at the second stage. As a result of exhaustion, increasing her risk of instrumental or caesarean deliveries. Mothers may become dissatisfied with service and may influence their health-seeking behaviour at subsequent births when deprived of meals and food.

Half of the women (51.4%) received labour analgesia with a third (29.4%) having episiotomy or laceration at childbirth. This number is significant especially when an episiotomy is not a recommended routine procedure, the experience of pain may affect the overall birthing experience for the women. The majority (80.2%) of the postnatal mothers had their babies with them within an hour of birth as a recommended practice by WHO, and mothers who were denied, may have their babies requiring additional support or critically ill after delivery [22]. The study revealed that women encouraged to have a support person, a woman interested with the idea of a birth companion, freedom of movement during labour, encouraged to eat during labour, labour analgesia and early bonding of mother and baby after delivery were significantly associated with the level of satisfaction of postnatal women towards the care received.

There was also a significant association on the forms of support received thus the woman encouraged to have a support person, liking the idea of a support person, the presence of labour companion, freedom of movement during labour, intake of fluids and meals labour analgesia and early bonding of mother and baby immediately after delivery with the experience of at least one form of disrespectful care. Therefore, a woman in labour denied movement, intake of fluids, meals, pain relief support person and early bonding of mother and baby has not received respectful maternity care.

### Experience of maltreatment during child birth

The majority (75.7%) of the women reporting for labour and delivery experienced at least a form of disrespectful care. This is similar to what was identified in studies done earlier in some parts of the country [13,15]. One in five of the women (19.8%) experienced physical abuse, about one in ten (9.3%) were discriminated against, about four in ten (38.9%) had undignified care, six in ten (60.4%) received unconsented care, about four in ten (34.5%) received non-confidential care, about one in five (17.9%) felt neglected and one in ten (7.7%) experienced detention against their will. The high prevalence of disrespectful care coupled with the representation of all the seven categories of disrespect and abuse in the study indicates that it is a problem requiring strengthening of the policy of respectful maternity care and institutional reforms. Failure to address this menace will cause clients to mistrust the healthcare system leading to poor health-seeking behaviour and the loss in battle against maternal morbidity and mortality.

There was a significant association between marital status, the highest level of education, employment, religion mode of delivery and health facility where childbirth occurred with the experience of at least one form of disrespectful care. The experience of at least one form of disrespectful care was 1.3times higher among single women (85.5%) than the married women (64.9%) and 1.4times higher among the divorced/widowed (92.3%) than the married. There was no significant association observed in the study of adolescent girls and women less than 25years with the experience of at least a form disrespectful care and hence contradicts with earlier studies of mistreatment among adolescent girls during childbirth [23,24].

## Conclusion

The study concludes that women received some forms of support during childbirth and labour but was marginally low especially in the area of partners presence during the birthing process. Most of the women experienced disrespectful care with all the categories represented. Expanding health facilities may not achieve desired skilled or facility-based delivery without improving the birth experience of women.

## Data Availability

All data that support the conclusions of the study are available in the manuscript

## Acknowledgement

The authors wish to thank all the women who participated in the study and providing frank response to the questionnaire.

## References

1. US Census Bureau. Measuring Maternal Mortality PRB. 2015; 1–6.

2. Floyd BL, Coulter N, Asamoah S, Agyare-asante R. Women ‘ s views and experience of their maternity care at a referral hospital in Ghana. 2015.

3. Bohren MA, Hunter EC, Munthe-kaas HM, Souza JP, Vogel JP. Facilitators and barriers to facility-based delivery in low- and middle-income countries : a qualitative evidence synthesis. 2014; 1,15. doi:10.1186/1742-4755-11-71

4. WHO. Strategies toward ending preventable maternal mortality (EPMM). Who. 2015;6736: 8. doi:ISBN 9789241508483

5. Souza J, Tunçalp Ö, Vogel J, Bohren M, Widmer M, Oladapo O, et al. Obstetric transition: the pathway towards ending preventable maternal deaths. BJOG An Int J Obstet Gynaecol. 2014;121: 1–4. doi:10.1111/1471-0528.12735

6. Rosen HE, Lynam PF, Carr C, Reis V, Ricca J, Bazant ES, et al. Direct observation of respectful maternity care in five countries: A cross-sectional study of health facilities in East and Southern Africa. BMC Pregnancy Childbirth. 2015;15: 10. doi:10.1186/s12884-015-0728-4

7. WHO. Harmonizing mind and body, people and systems People at the Centre of Health Care. 20 Avenue Appia, 1211 Geneva 27, Switzerland; 2007.

8. Nair M, Yoshida S, Lambrechts T, Boschi-Pinto C, Bose K, Mason EM, et al. Facilitators and barriers to quality of care in maternal, newborn and child health: a global situational analysis through metareview. BMJ Open. 2014;4: e004749. doi:10.1136/bmjopen-2013-004749

9. Knight HE, Self A, Kennedy SH. Why Are Women Dying When They Reach Hospital on Time? A Systematic Review of the ‘Third Delay.’ Young RC, editor. PLoS One. 2013;8: e63846. doi:10.1371/journal.pone.0063846

10. Windau-Melmer T. A Guide for Respectful Maternity Care. Washington DC; 2013.

11. White Ribbon Alliance. Respectful Maternity Care Charter. 2011.

12. Sando D, Ratcliffe H, McDonald K, Spiegelman D, Lyatuu G, Mwanyika-Sando M, et al. The prevalence of disrespect and abuse during facility-based childbirth in urban Tanzania. BMC Pregnancy Childbirth. 2016;16. doi:10.1186/s12884-016-1019-4

13. Asefa A, Bekele D. Status of respectful and non-abusive care during facility-based childbirth in a hospital and health centers in Addis Ababa, Ethiopia. Reprod Health. 2015;12: 7,8. doi:10.1186/s12978-015-0024-9

14. Okafor II, Ugwu EO, Obi SN. Disrespect and abuse during facility-based childbirth in a low-income country. International Journal of Gynecology and Obstetrics. 2014. pp. 110–113. doi:10.1016/j.ijgo.2014.08.015

15. Moyer CA, Managing MPH, Adongo PB, Lecturer S, Aborigo RA, Health MPH, et al. ‘ They treat you like you are not a human being ‘ : Maltreatment during labour and delivery in rural northern Ghana. Midwifery. 2013; 4,7. doi:10.1016/j.midw.2013.05.006

16. Ghana Statistical Service. Ghana demographic and health survey 2014. Ghana demographic and health survey 2014. 2014. doi:10.1007/b138909

17. Siraj A, Teka W, Hebo H. Prevalence of disrespect and abuse during facility based child birth and associated factors, Jimma University Medical Center, Southwest Ethiopia. BMC Pregnancy Childbirth. 2019;19: 185. doi:10.1186/s12884-019-2332-5

18. Moyer CA, Rominski S, Nakua EK, Dzomeku VM, Agyei-Baffour P, Lori JR. Exposure to disrespectful patient care during training: Data from midwifery students at 15 midwifery schools in Ghana. Midwifery. 2016;41: 39–44. doi:10.1016/j.midw.2016.07.009

19. Reis V, Deller B, Senior M, Advisor M, Carr C. Respectful Maternity Care Country experiences Survey Report. 2012.

20. Mensah RS, Mogale RS, Richter MS. Birthing experiences of Ghanaian women in 37th Military Hospital, Accra, Ghana. Int J Africa Nurs Sci. 2014;1: 29–34. doi:10.1016/j.ijans.2014.06.001

21. Perkins J, Ehsanur Rahman A, Mhajabin S, Bakkar Siddique A, Mazumder T, Rifat Haider M, et al. Sexual and Reproductive Health Matters. 2019;27: 228–247. doi:10.1080/26410397.2019.1610277

22. World Health Organization (WHO). Intrapartum care for a positive childbirth experience WHO recommendations. 2018.

23. Bowser D, Hill MPHK. Exploring Evidence for Disrespect and Abuse in Facility-Based Childbirth Report of a Landscape Analysis. 2010.

24. Maya ET, Adu-Bonsaffoh K, Dako-Gyeke P, Badzi C, Vogel JP, Bohren MA, et al. Women’s perspectives of mistreatment during childbirth at health facilities in Ghana: findings from a qualitative study. Reprod Health Matters. 2018;26. doi:10.1080/09688080.2018.1502020

